# The Efficacy and Safety of Daily Low-Dose Iron Supplementation in Adults with Dietary Iron Deficiency: a Double-Blind, Randomized, Single-Center Study

**DOI:** 10.64898/2026.02.24.26346946

**Authors:** Andrej Kravos, Blaž Dolenc, Nana Fartek, Igor Locatelli, Nanča Čebron Lipovec, Neža Rogelj Meljo, Mitja Kos, Tisa Dobovšek, Gabriela Panter

## Abstract

Iron deficiency (ID) is the most common nutritional deficiency worldwide, often caused by insufficient dietary intakes. Oral supplementation is one of the means to improve iron status. This study evaluated the efficacy and safety of two low-dose iron supplements - *>Your< Iron Forte Capsules (YIFC)* and *Ferrous Sulfate Capsules (FSC)* - in individuals with dietary ID. One hundred and one participants (mean age 30.6 years; 98% women) with low iron stores (mean serum ferritin 16.1 µg/L) were randomized to receive either YIFC or FSC once daily for 12 weeks. Changes in blood indices and iron-related parameters were assessed at four and 12 weeks of intervention relative to baseline. The primary outcome was the change in hemoglobin (Hb) after 12 weeks. Eighty-seven participants completed the study. Both supplements significantly increased Hb at 12 weeks (YIFC: mean 6.52 g/L, p<0.001; FSC: mean 5.71 g/L, p<0.001). Product-related adverse events (AEs) were few (17% of all AEs) and of mild to moderate intensity only. One participant receiving FSC withdrew due to a probable product-related AE. The frequencies of product-related AEs were similar between study arms, however, statistically significantly more AEs judged to be definitely related to the product occurred in in the FSC arm. While product-related AEs were confined to the gastrointestinal tract in the YIFC arm, they affected multiple organ systems in the FSC arm. Supplementation with either YIFC or FSC proved as an effective, well-tolerated, and safe strategy for improving iron status in non-anemic dietary iron deficiency. In terms of the AE profile, supplementation with YIFC may offer advantages over supplementation with FSC.

## Introduction

Iron deficiency (ID) is the most prevalent nutritional deficiency worldwide and is a leading cause of anemia ^1–3^. Iron deficiency, with or without accompanying anemia, may manifest as an isolated condition caused by inadequate dietary intake, or as a comorbidity linked to blood loss or malabsorption due to gastrointestinal disorders, medication use, infections, or chronic non-communicable diseases. The etiology of and contributing factors to ID prevalence are influenced by a complex interplay of demographic, geographic, and environmental variables ^2–4^.

Specific population groups, notably children, adolescents, and premenopausal or pregnant women, are at a particularly high risk for the development of ID due to elevated physiological iron demands ^3^, often unmet by dietary iron intakes. Studies on dietary iron intake among European women have revealed inadequacy rates ranging from 61% to 97%, corresponding to ID prevalence estimates of 10% to 40% ^5–7^. Preventing ID is a recognized public health priority for women due to its far-reaching implications for overall health, well-being, productivity and quality of life - maintaining an adequate iron status is critical for preserving physical, cognitive, and immune performance and for supporting maternal and child health ^2,4,8^.

Among various etiologies of ID, dietary ID is the most straightforward to address, as it does not require the management of underlying medical conditions beyond nutritional insufficiency. Accordingly, dietary intake constitutes the most critical modifiable determinant of iron status. Food-based strategies to increase dietary intake of iron include dietary modification and diversification, iron supplementation, and food fortification ^3^. While fortification serves as a population-level intervention, dietary change and supplementation offer individualization, allowing for tailored nutritional management.

Long-term adherence to strategies for maintaining adequate iron status is generally required, particularly for individuals at risk of ID, and while dietary modification and diversification are generally considered first-line interventions for early-stage, non-anemic ID, real-world adherence to these strategies is often suboptimal, as many individuals encounter difficulties in consistently altering the type, quantity, or timing of food intake. For example, 80% of participants following a daily dietary regimen with a beverage high in vitamin C content to aid in absorption of iron declared that they would not continue to consume the beverage with the same frequency beyond the study, and more than half of participants in another study of dietary intervention vs. supplementation found it difficult to consume the required amounts of iron-containing foods, resulting in mean compliance with the dietary regime of only 58% ^9,10^. On the other hand, daily supplementation was adhered to by more than 90% of participants and was stated to be a preferred option of intervention by 62% of participants, because it was found easier ^9^. Therefore, supplementation may offer a more effective and feasible alternative for addressing dietary ID in practice, especially as it has gained prominence as a common lifestyle behavior in industrialized nations, where up to 60% of individuals report regular use of dietary supplements and demonstrate relatively high long-term adherence rates ^11–13^. This trend suggests that iron supplementation, when appropriately indicated, may provide a more sustainable and impactful intervention for preventing and managing ID by bridging the gaps between actual and optimal iron intakes.

Despite the widespread availability of iron supplements on the market, most products lack rigorous scientific and clinical validation, particularly in iron deficient, yet non-anemic populations, limiting the ability of healthcare providers and consumers to make informed, evidence-based decisions regarding their efficacy and safety in non-patient populations. Furthermore, direct comparisons with gold-standard iron formulations, such as ferrous sulfate, are generally absent. Many supplements also deliver iron in relatively high doses, often exceeding tolerable intake thresholds, resulting in gastrointestinal side effects commonly associated with therapeutic-dose iron supplementation. In light of these challenges, the present study aimed to provide robust clinical substantiation of a novel Qfer® ingredient-based low-dose iron-containing dietary supplement, *>Your< Iron Forte Capsules (YIFC)*, and a well-established ferrous sulfate-based supplement, *Ferrous Sulfate Capsules (FSC)*, that has through dose reduction been repurposed from therapeutic to supplemental use. We hypothesized that both supplements would effectively improve overall iron status and demonstrate similarly favorable tolerability profiles, contributing to high acceptability among participants. Additionally, based on prior research on ID in developed countries, we anticipated a relatively high prevalence of dietary ID within the population, particularly among women, underscoring the broader relevance of this research, contributing to ongoing discussions on women’s health and nutrition policies, and highlighting the need for targeted dietary interventions to prevent ID and support overall health and well-being.

## Results

### Participants

One hundred and one individuals (99 women and two men) were randomized to an intervention. Arm allocation and progression of participants through the study is summarized in **Figure 1**.

**Figure 1.**
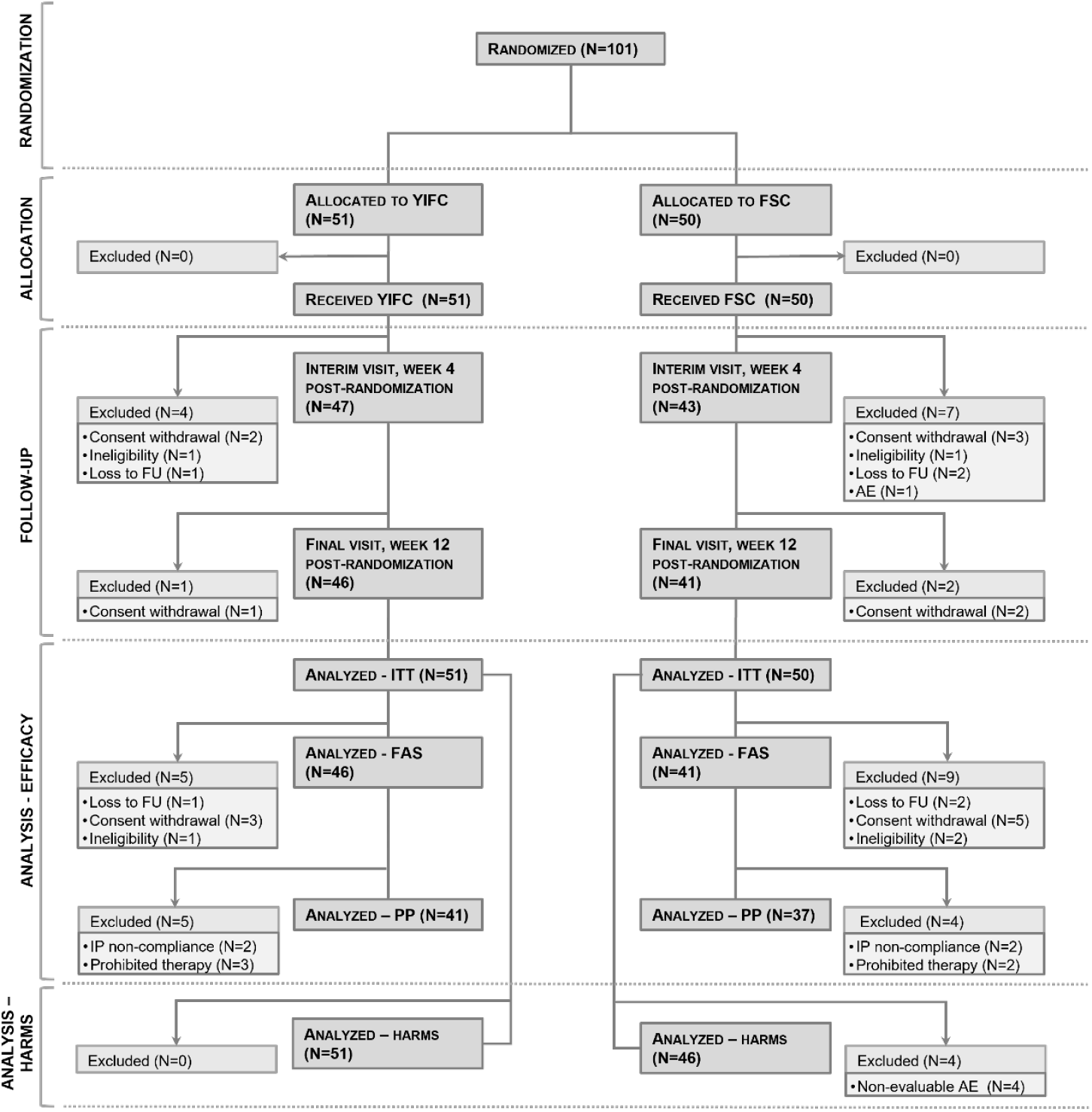
Participant flow and populations for analyses. YIFC: >Your< Iron Forte Capsules, FSC: Ferrous Sulfate Capsules, FU: follow-up, AE: adverse event, IP: investigational product, ITT: intention-to-treat, FAS: full analysis set, PP: per protocol

The total attrition rate after randomization was 14%. Eleven participants (11%) dropped out by or at their second study visit: five withdrew consent, three were lost to follow-up, one was excluded due to experiencing an allergic skin reaction judged to be probably related to the consumption of the IP, and two became ineligible - one began receiving exclusionary therapy, and the other one was scheduled for major surgery due to a deterioration of an underlying medical condition. An additional three participants (3%) dropped out between the second and third visits or at the time of the third visit - all three withdrew consent. Hence, 87 individuals (85 women and two men) completed the study and were included in the FAS for efficacy analyses.

### Participant Baseline Demographic and Clinical Characteristics

Participant baseline characteristics were well balanced between the two study arms with no statistically significant differences in any variables (**Table 1**). The population consisted almost entirely of women (98%), aged 18–49 years, with a healthy BMI. Forty-four women (35%) reported a total of 91 previous pregnancies and 83 births. Additionally, 57% of participants were regular users of dietary supplements. Hematological profiles were indicative of early-stage iron depletion without overt signs of impaired erythropoiesis: the level of storage iron was low (mean SF level 16 µg/L), but values of transport (serum iron, TIBC) and functional (Hb, RBCs) iron indices remained within normal population reference ranges.

**Table 1.** Baseline demographic and clinical characteristics of study participants. Data is shown for the FAS population.

| Parameter | Study population*<br>by intervention arm |  | ALL (N=87) |
| --- | --- | --- | --- |
|  | YIFC (N=46) | FSC (N=41) |  |
| Demographics |  |  |  |
| Age (years), mean (SD) | 31.0 (9.1) | 30.1 (8.1) | 30.6 (8.6) |
| Gender, N (%) of females | 45 (98) | 40 (98) | 85 (98) |
| Weight (kg), mean (SD) | 63.3 (7.2) | 60.6 (8.2) | 62.1 (7.7) |
| BMI (kg/m2), mean (SD) | 22.0 (2.6) | 21.5 (2.1) | 21.8 (2.4) |
| Important medical history, N (%) | 7 (15) | 6 (15) | 13 (15) |
| Concomitant medical conditions, N (%) | 4 (9) | 4 (10) | 8 (9) |
| Regular dietary supplement use, N (%) | 26 (57) | 24 (59) | 50 (57) |
| RBC indices |  |  |  |
| Erythrocytes (10<sup>12</sup>/L), mean (SD) | 4.3 (0.3) | 4.3 (0.3) | 4.3 (0.3) |
| Hb (g/L), mean (SD) | 121.2 (9.4) | 122.7 (9.1) | 121.9 (9.2) |
| Hct, mean (SD) | 0.365 (0.028) | 0.368 (0.025) | 0.366 (0.026) |
| MCV (fL), mean (SD) | 85.0 (5.6) | 84.9 (4.2) | 85.0 (5.0) |
| MCH (pg), mean (SD) | 28.2 (2.4) | 28.3 (1.8) | 28.2 (2.2) |
| MCHC (g/L), mean (SD) | 331.1 (10.7) | 332.9 (9.1) | 332.8 (12.0) |
| Iron panel |  |  |  |
| Serum iron (μmol/L), mean (SD) | 11.2 (6.8) | 13.0 (6.4) | 12.0 (6.6) |
| TIBC (μmol/L), mean (SD) | 58.2 (7.7) | 60.0 (7.6) | 59.1 (7.6) |
| TSAT (%), mean (SD) | 19.8 (12.6) | 22.1 (11.7) | 20.9 (12.2) |
| SF (μg/L), mean (SD) | 16.0 (6.8) | 16.1 (7.0) | 16.1 (6.9) |

### Follow-up Hematological Characteristics and Iron Status

The efficacy results for the primary and secondary outcomes are presented in **Tables 2 and 3**, respectively. For the primary outcome, a change in Hb, significant improvements were demonstrated at both four and 12 weeks into the intervention period (**Table 2**).

**Table 2.** Change in hemoglobin by baseline hemoglobin status and study intervention arm. Data is shown for the FAS population. Hb: hemoglobin, Overall baseline Hb: low and normal baseline Hb combined, Low baseline Hb: <120 g/L for women and <130 g/L for men, Normal baseline Hb: ≥120 g/L for women and ≥130 g/L for men, YIFC: >Your< Iron Forte Capsules, FSC: Ferrous Sulfate Capsules.

| Intervention arm | Hb<br>mean (SD) |  |  | Absolute change in Hb<br>mean (SD), (95% CI for mean), p-value |  |
| --- | --- | --- | --- | --- | --- |
|  | baseline | 4 weeks | 12 weeks | 4 weeks vs. baseline | 12 weeks vs. baseline |
| **Overall baseline Hb** |  |  |  |  |  |
| YIFC (N=46) | 121.2 (9.4) | 123.7 (9.7) | 127.7 (9.4) | 2.46 (5.70), (0.76-4.15), **p=0.005** | 6.52 (7.14), (4.40-8.64), **p<0.001** |
| FSC (N=41) | 122.7 (9.1) | 124.7 (7.5) | 128.4 (7.8) | 2.07 (5.81), (0.24-3.91), **p=0.028** | 5.71 (9.10), (2.83-8.58), **p<0.001** |
| **Low baseline Hb** |  |  |  |  |  |
| YIFC (N=21) | 114.43 (6.89) | 118.38 (9.74) | 124.76 (9.85) | 3.95 (5.97), (1.23-6.67), **p=0.007** | 10.33 (7.19), (7.06-13.60), **p<0.001** |
| FSC (N=13) | 112.08 (4.65) | 118.69 (5.38) | 124.69 (7.83) | 6.62 (6.95), (2.42-10.81), **p=0.005** | 12.63 (11.12), (5.90-19.33), **p=0.001** |
| **Normal baseline Hb** |  |  |  |  |  |
| YIFC (N=25) | 126.88 (7.27) | 128.08 (7.25) | 130.20 (8.41) | 1.20 (5.25), (-0.97-3.37), p=0.265 | 3.32 (5.39), (1.09-5.55), **p=0.005** |
| FSC (N=28) | 127.57 (5.78) | 127.54 (6.74) | 130.07 (7.25) | -0.04 (3.74), (-1.48-1.41), p=0.960 | 2.50 (5.85), (0.23-4.77), **p=0.032** |

**Table 3.** Summary of efficacy outcomes by study intervention arm. Data is shown for the FAS population. YIFC: >Your< Iron Forte Capsules, FSC: Ferrous Sulfate Capsules, RBC: red blood cell, BMI: body mass index, Hct: hematocrit, MCV: mean corpuscular volume, MCH: mean corpuscular hemoglobin, MCHC: mean corpuscular hemoglobin concentration, TSAT: transferrin saturation, SF: serum ferritin.

| Intervention arm | Outcome mean (SD) |  |  | Absolute change in outcome mean (SD), (95% CI for mean), p-value |  |
| --- | --- | --- | --- | --- | --- |
|  | baseline | 4 weeks | 12 weeks | 4 weeks vs. baseline | 12 weeks vs. baseline |
| **Hct** |  |  |  |  |  |
| YIFC (N=46) | 0.365 (0.028) | 0.373 (0.025) | 0.382 (0.026) | 0.008 (0.024), (0.001-0.016), **p=0.020** | 0.017 (0.025), (0.010-0.025), **p<0.001** |
| FSC (N=41) | 0.368 (0.025) | 0.376 (0.020) | 0.380 (0.021) | 0.008 (0.021), (0.001-0.014), **p=0.024** | 0.012 (0.025), (0.003-0.020), **p=0.006** |
| **MCV (fL)** |  |  |  |  |  |
| YIFC (N=46) | 85.0 (5.6) | 85.5 (5.3) | 86.3 (4.4) | 0.52 (1.18), (0.16-0.87), **p=0.005** | 1.27 (2.49), (0.54-2.01), **p=0.001** |
| FSC (N=41) | 84.9 (4.2) | 86.2 (3.5) | 86.9 (3.1) | 1.24 (1.71), (0.70-1.78), **p<0.001** | 1.97 (2.78), (1.09-2.85), **p<0.001** |
| **MCH (pg)** |  |  |  |  |  |
| YIFC (N=46) | 28.2 (2.4) | 28.4 (2.4) | 28.9 (2.1) | 0.18 (0.44), (0.05-0.31), **p=0.008** | 0.70 (0.78), (0.46-0.93), **p<0.001** |
| FSC (N=41) | 28.3 (1.8) | 28.8 (1.7) | 29.4 (1.3) | 0.46 (0.46), (0.31-0.60), **p<0.001** | 1.07 (1.08), (0.73 - 1.41), **p<0.001** |
| **MCHC (g/L)** |  |  |  |  |  |
| YIFC (N=46) | 331.1 (10.7) | 331.1 (10.0) | 334.5 (10.5) | -0.02 (5.39), (-1.62-1.58), **p=0.978** | 3.35 (9.79), (0.44-6.26), **p=0.025** |
| FSC (N=41) | 332.9 (9.1) | 333.5 (10.6) | 338.0 (9.1) | 0.59 (6.00), (-1.31-2.48), **p=0.536** | 5.12 (8.35), (2.49-7.76), **p<0.001** |
| **TSAT (%)** |  |  |  |  |  |
| YIFC (N=46) | 19.8 (12.6) | 28.0 (16.2) | 28.5 (13.5) | 8.24 (17.82), (2.95-13.53), **p=0.003** | 8.72 (17.42), (3.54-13.89), **p=0.001** |
| FSC (N=41) | 22.1 (11.7) | 29.7 (15.7) | 31.0 (15.9) | 7.61 (16.66), (2.35-12.87), **p=0.006** | 8.88 (16.54), (3.66-14.10), **p=0.001** |
| **SF (µg/L)** |  |  |  |  |  |
| YIFC (N=46) | 16.0 (6.8) | 22.5 (10.4) | 25.3 (10.6) | 6.41 (9.03), (3.73-9.09), **p<0.001** | 9.30 (9.87), (6.37-12.24), **p<0.001** |
| FSC (N=41) | 16.1 (7.0) | 31.1 (15.9) | 36.4 (18.4) | 14.96 (13.56), (10.68-19.24), **p<0.001** | 20.27 (17.14), (14.86-25.68), **p<0.001** |

A subgroup analysis, performed by stratifying participants according to their baseline Hb status, revealed that individuals with low baseline Hb exhibited more pronounced improvements in Hb at both four and 12 weeks of intervention as compared to those with normal baseline Hb. When compared, the difference in Hb values after 12 weeks of intervention between the YIFC and the FSC arms were not statistically significant in either of the Hb-stratified groups (p=0.642 for overall baseline Hb, p=0.471 for low baseline Hb, p=0.599 for normal baseline Hb), however, the study was not designed to detect a between-arm difference. A separate non-inferiority analysis using a 4 g/L margin on the overall baseline Hb population demonstrated that YIFC were statistically significantly non-inferior to FSC in increasing Hb levels (p=0.001). For other secondary outcomes, the dynamics of improvements in SF, TSAT and RBC indices in YIFC and FSC arms paralleled those of Hb, showing statistically significant improvements from baseline at both four and 12 weeks of intervention (**Table 3**).

The only RBC index failing to reach statistical significance at four weeks of intervention at four weeks of intervention was MCHC, however, in ID, RBCs will preserve Hb at the expense of RBC size and MCHC will consequently be the last RBC index to decrease. Consequently, in a population with latent ID, where all RBC indices still reside within reference ranges, increments upon iron supplementation are expected to be modest, especially in indices least affected by ID. While SF reflects iron stores, it is also a positive acute-phase reactant and can be elevated in the presence of inflammation, potentially leading to an overestimation of iron stores. To account for this, CRP was measured alongside SF as a marker of inflammation. Elevated CRP levels (>10 mg/L) were observed in one participant in the FSC arm at the week four post-randomization assessment and in two participants in the same arm at week 12 post-randomization. Excluding these data points from the analysis did not affect the study outcomes. Similarly, excluding data on male participants to evaluate outcomes in an all-female population also did not result in meaningful changes to the results.

### Compliance with Assigned Interventions

Compliance with iron-containing products is one of the crucial factors in determining the success of oral iron repletion. Compliance with IPs throughout the study is presented in **Table 4**. Overall, 99% of participants adhered to the dosing schedule throughout the study period. Compliance with IPs was excellent, with no significant differences between four and 12 weeks of intervention.

**Table 4.**
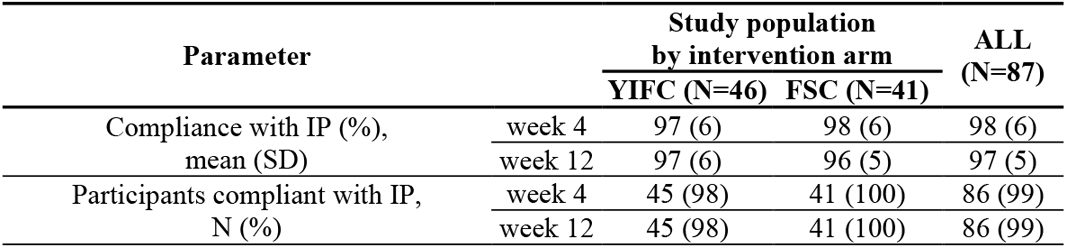
Compliance with investigational product consumption by study intervention arm and study investigational product. Data is shown for the FAS population.

### Adverse Events

Harms were recorded for each participant from the time of informed consent until their final day of study participation. Of the 101 randomized participants, 16 reported no AEs, while the remaining 85 participants reported altogether 334 AEs. Of these, 330 AEs experienced by 81 participants could be fully characterized and were analyzed in detail, as presented in **Supplementary Table 1**. Of the 330 evaluable AEs, altogether 56 Aes occurring in 36 participants were considered possibly, probably, or definitely related to IP. Details on IP-related AEs are presented in **Table 5**.

**Table 5.**
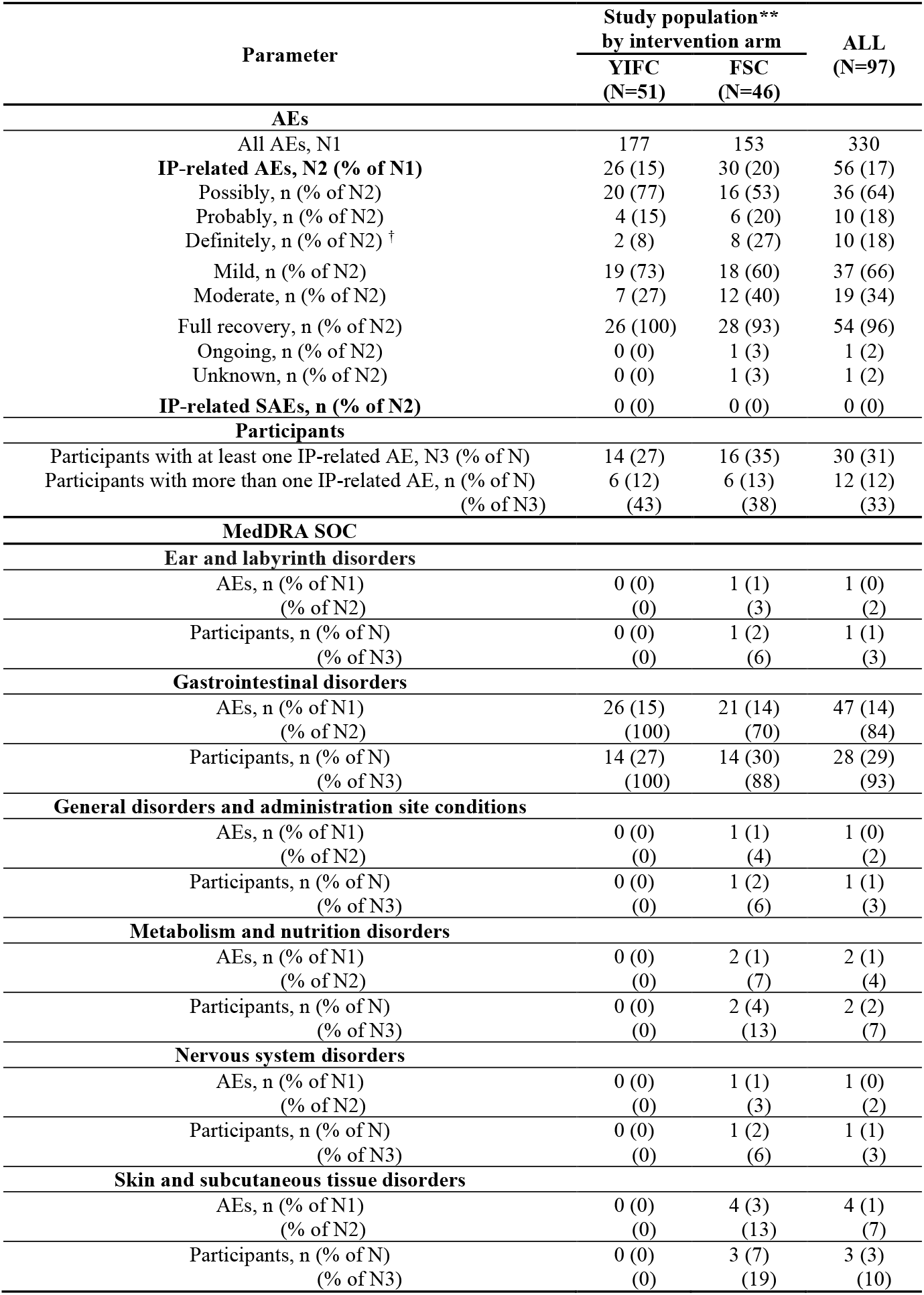
Summary of investigational product-related* adverse events by study intervention arm. * Investigational product-related IPs: those judged to be possibly, probably, or definitely related. ** Safety population. YIFC: >Your< Iron Forte Capsules, FSC: Ferrous Sulfate Capsules, AE: adverse event, MedDRA: medical dictionary for regulatory activities, SOC: system organ class. ^†^ Statistically significantly different at p<0.05.

**Table 6.** Summary of changes in energy/fatigue by intervention arm. Results presented are for the FAS population. N_0:_ number of evaluable participants at baseline, N_4:_ number of evaluable participants at four weeks into the intervention, N_12:_ number of evaluable participants at 12 weeks into the intervention. The number of evaluations for each arm and each timepoint differ from the number of individuals in the entire FAS population due to missing data.

| Intervention arm | Outcome<br>mean (SD) |  |  | Absolute change in outcome<br>mean (SD), (95% CI for mean), p-value |  |
| --- | --- | --- | --- | --- | --- |
|  | baseline | 4 weeks | 12 weeks | 4 weeks vs. baseline | 12 weeks vs. baseline |
| **Energy/Fatigue** |  |  |  |  |  |
| YIFC (N<sub>0</sub>=42, N<sub>4</sub>=42, N<sub>12</sub>=40) | 53.6 (18.9) | 64.9 (17.8) | 67.1 (16.0) | 11.3 (16.5), (6.2-16.5), **p<0.001** | 14.0 (15.3), (9.1-18.9), **p<0.001** |
| FSC (N<sub>0</sub>=40, N<sub>4</sub>=40, N<sub>12</sub>=40) | 50.3 (21.9) | 58.6 (19.6) | 64.8 (18.0) | 8.4 (20.6), (1.8-15.0), **p=0.014** | 14.5 (20.6), (7.9-21.1), **p<0.001** |

None of the IP-related AEs met the criteria for a serious adverse event (SAE) and all were of either mild (37 (66%) AEs) or moderate (19 (34%) AEs) intensity. Ninety-six percent of IP-related AEs were of a transient nature with 93% of participants achieving a full recovery. One gastrointestinal AE remained ongoing at study completion, and the status of another IP-related AE in a different participant was unknown due to loss of follow-up. There were no significant differences in the number of participants experiencing IP-related AEs or the frequency of IP-related AEs between the study arms, with the exception of AEs classified as definitely related to the IP, which occurred significantly less often in the YIFC arm. Most IP-related AEs were classified under the Gastrointestinal disorders MedDRA SOC. IP-related AEs classified into SOCs other than Gastrointestinal disorders occurred exclusively in the FSC arm and were dispersed through the Ear and labyrinth disorders (one AE (vertigo)), General disorders and administration site conditions (one AE (MedDRA Preferred Term (PT) malaise)), Metabolism and nutrition disorders (two AEs in two participants (MedDRA PTs appetite disorder and increased appetite), Nervous system disorders (one AE (MedDRA PT headache)), and Skin and subcutaneous tissue disorders (four AEs in three participants (MedDRA PTs atopic dermatitis, rash, acne, and rosacea)).

### Energy and Fatigue

Participant self-assessments of energy/fatigue over the preceding four weeks were performed at baseline and both four and 12 weeks of intervention using the vitality subscale of the RAND-SF36 questionnaire. Changes in energy/fatigue over time are presented in **Table 7**. Significant improvements from baseline were observed in both study arms, however, when using the minimal clinically important difference (MCID) defined as half a standard deviation (SD) ^14^, the YIFC exceeded the MCID threshold at both four and 12 weeks, whereas FSC only did so at 12 weeks.

## Discussion

This study aimed to evaluate the potential of low-dose iron dietary supplementation to improve iron status among adults in a developed country presenting with dietary ID. Due to the absence of routine public screening programmes for detecting ID in healthy, at-risk populations, eligible participants were identified through study-specific mass screening of volunteers. Although both sexes were targeted during recruitment, the final study population consisted almost entirely of women (98%). This outcome was anticipated, as women of childbearing age are considered an at-risk population for developing ID, with the primary contributing factors being repetitive iron loss through menstruation and increased iron requirements during pregnancy, both often accompanied by insufficient dietary iron intakes ^2,8^. While we did not collect or analyze dietary data to confirm a nutritional cause of ID in our participants, employing strict inclusion and exclusion criteria and rigorous screening procedures at early stages of eligibility assessment allowed for selective exclusion of individuals in whom ID might have been secondary to non-dietary causes (e. g., malabsorption, gastrointestinal bleeding, certain medications). By using this approach and applying a criterion of SF<30 µg/L as an indicator of depleted iron stores, we identified dietary ID in 27% of screened women and 5% of screened men. These figures align well with recent data from a representative sample of the Slovenian population, which reported inadequate daily iron intake in 76% of premenopausal women and 9% of men, resulting in SF <30 µg/L in 27% of women and 4% of men ^7^. We identified 13% of screened women to have SF levels even below 15 µg/L, which, according to World Health Organization (WHO) criteria, can already be interpreted as a public health concern ^15^.

Collectively, these findings challenge the common assumption that dietary needs are consistently met in middle- and high-income settings due to overall food abundance and underscore the rationale for implementing targeted screening programs for the early detection of ID as well as the potential benefit of sustained supplementation regimes in at-risk population groups even in developed countries.

Both YIFC and FSC effectively improved the iron status in our iron-deficient population, exhibiting similar iron-repletion profiles, with the magnitude and the dynamics of changes within physiological levels and consistent with the population baseline iron status, the dose of iron provided, and the duration of the intervention. Iron deficiency in the study population was characterized by depleted iron stores without clinical signs of overt anemia. Hemoglobin concentrations, RBC indices, and iron metabolism-related parameters remained within normal reference ranges, though values tended to cluster near the lower limits, suggesting a mild impairment in hematologic function. This is consistent with existing evidence indicating that signs of iron-deficient erythropoiesis can begin to emerge at SF levels below 40 µg/L ^16^. While low iron stores alone, even in the absence of effects on transport and functional iron, suffice to suppress hepcidin synthesis and thereby enhance iron absorption, these regulatory responses are more pronounced in advanced ID and particularly in iron deficiency anemia (IDA), where erythropoiesis is significantly impaired and Hb levels therefore markedly reduced [4,24]. As a result, individuals with IDA typically exhibit greater increases in Hb concentrations following supplementation compared to those with non-anemic ID ^17,18^. This pattern was also evident in our study; a subgroup analysis based on baseline Hb levels showed that participants with lower baseline Hb (<120 g/L) experienced a more pronounced increase in Hb following iron supplementation compared to those with higher baseline Hb (≥120 g/L). While a rise of 10–20 g/L in Hb over three to four weeks is considered an adequate response in IDA, such an increase clearly cannot be expected to occur in non-anemic ID, as it would be non-physiological ^4^. Instead, mean improvements in Hb concentration of 6–7 g/L as observed in our study can be considered appropriate for non-anemic ID on a low daily dose of iron and are also concordant with average increases of 3 g/L–6 g/L reported in other studies of iron supplementation in non-anemic iron-deficient menstruating women ^17,19–21^. By the end of the intervention, mean Hb concentrations in both study arms were well within the normal ranges, and it is unlikely that any further significant increases in Hb concentration would have been achieved by prolonging the duration of the intervention.

In contrast to Hb, SF levels would likely have continued to improve with sustained supplementation. Although both intervention groups showed substantial increases in SF over 12 weeks, only the ferrous sulfate group achieved a mean SF above 30 µg/L. Even so, 40% of participants in this group remained iron-deficient at the end of the study. This is not unexpected, as iron repletion is a gradual process. In cases of IDA, once Hb levels are corrected, it can take an additional three to six months or even up to a year of continued supplementation to fully restore iron stores ^1,22^. On high-dose iron therapy, SF typically increases by 2–5 µg/L per week, with stores generally replenished within 6–12 months ^4,22^. However, with low-dose supplementation, both the rate and magnitude of SF increase are lower, and reaching a steady state takes longer ^17,18^. This can be attributed to differences in iron absorption: while lower doses of ferrous sulfate (e. g., 40 mg) exhibit higher fractional absorption than higher doses (>60 mg) in iron-depleted individuals, the total amount of iron absorbed remains greater with therapeutic doses ^23,24^.

Therapeutic doses of iron, while effective, present clear limitations, particularly in the context of dietary interventions aimed at addressing inadequate iron intake in otherwise healthy individuals. High-dose iron supplementation is frequently associated with gastrointestinal side effects such as nausea, vomiting, constipation, diarrhea, and abdominal discomfort, which are often severe enough to significantly reduce compliance and therefore compromise the efficacy and real-world effectiveness of interventions ^25,26^. In our study, compliance was excellent, being at or above 97% at 12 weeks of intervention, with only one participant discontinuing due to side effects. This suggests good acceptance of both the dosing regimen and its tolerability within the controlled study environment.

Furthermore, the interventions used in this study appear well-suited for effective use in real-world settings; as dietary supplements rather than medications, the investigational products used in this study offer two key advantages: greater user acceptability and ease of integration into daily routines. The low iron content in the daily dose significantly reduces the likelihood of adverse effects ^17^ and in our study, both supplements were indeed well tolerated, with 74% of participants reporting no side effects, suggesting a high potential for continued use beyond the study context. Additionally, the use of dietary supplements to support general health and well-being is increasingly common and socially accepted. In our sample, 74% of participants identified as dietary supplement users, and 58% reported regular use. These figures align with broader trends observed in other populations ^11,12^, indicating that supplementation is already a familiar practice for many. Given this context, the daily supplementation regimen used in our study can reasonably be expected to integrate smoothly into the existing lifestyles of most individuals in industrialized countries, requiring little to no modification of dietary habits.

This study has several strengths, but also some limitations that highlight the need for larger, placebo-controlled trials to replicate the findings and expand understanding by addressing additional factors relevant to iron metabolism and repletion. Although conducted at a single research center, participants were recruited from the broader metropolitan and urban region of central Slovenia. This approach yielded a more diverse study population than would have been possible if recruitment had been limited to a single healthcare facility or practitioner. While the study population may be considered broadly reflective of a generally healthy Western demographic, particularly women of reproductive age, who are most commonly affected by dietary ID due to inadequate intake relative to physiological needs, more detailed profiling of participants could have provided further insight into the underlying nature of dietary ID. For instance, ID may result from insufficient intake of iron or from its low bioavailability despite adequate intake, such as it occurs in vegetarian or vegan diets. Similarly, while 23% of female participants reported experiencing heavy menstrual bleeding, a known contributor to ID in premenopausal women, this was not quantitatively assessed to estimate its impact on iron status. Despite these limitations, the study population remains highly representative of the real-world target group for dietary iron interventions and the findings offer practical and clinically relevant insights into the characteristics of low-dose iron supplementation for improving iron status in otherwise healthy individuals. The relatively short duration of the study represents a notable limitation. Extending the intervention period could have provided valuable insights into both the long-term efficacy and the dynamics of iron store repletion. It would also have allowed for a more thorough evaluation of sustained compliance, which is critical for achieving full repletion and for assessing the feasibility of long-term or continuous supplementation.

Compliance and dropout rates in clinical trials may not fully reflect real-world adherence; discontinuation rates due to side effects in interventional studies typically range from 0% to 24% but can be as high as 40% in observational and population-based studies ^27^. For instance, in one trial using high-dose iron, 76% of participants reported adverse effects, yet only one person withdrew. Despite a reported mean compliance rate of 93%, over one-third of participants acknowledged that side effects interfered with their ability to take the supplement consistently. This highlights the likelihood that, outside a monitored study setting, compliance would be significantly lower, ultimately limiting the intervention’s effectiveness ^9^.

While the sample size was sufficient to detect within-group changes, a larger cohort would be necessary to reliably assess differences between the two intervention arms. Also, the study would have significantly benefited from the inclusion of a placebo arm in the study design. While placebo responses are unlikely to produce significant changes in objective, blood-based biomarkers of iron status, including a placebo group would have strengthened the validity of our conclusions by controlling for potential confounding factors, one such being participant behavior. Although instructed not to change their dietary or lifestyle habits during the study, the participants might have nevertheless done so; in response to learning about their ID, they might have, albeit inadvertently, adopted more favorable dietary and lifestyle choices affecting their overall iron status. Such changes could have independently influenced iron status and confounded the observed effects of supplementation.

Similarly, participant self-report outcomes of energy/fatigue could have been better evaluated in comparison with placebo, especially because of their subjective nature, making them more prone to contextual effects of an intervention. Fatigue as a hallmark symptom of ID, being reported by more than 90% of affected women, has been shown to improve upon iron supplementation, however, substantial placebo responses have been noted in some studies ^28–30^. Moreover, the assessment of fatigue using a symptom-specific participant-reported outcome measure could have been more informative than deriving the changes from the RAND-SF-36 questionnaire fatigue subscale. Therefore, findings related to fatigue in this study should be interpreted with caution and within the limited context.

A notable strength of this study is the use of iron-containing dietary supplements rather than pharmaceutical iron preparations. This aligns with current recommendations that advocate for dietary interventions as a first-line approach in managing non-urgent, non-anemic ID, aiming to nutritionally support individuals with inadequate dietary iron intake. The chosen dose and dosing schedule further reinforce this strength. Specifically, the daily provision of ≤40 mg of elemental iron is consistent with evidence supporting this regimen as optimal for maximizing fractional iron absorption in individuals with ID or mild IDA ^26^. Additionally, the daily supplementation of 30 mg of iron, as provided by the products tested in this study, complements the estimated mean intake of dietary iron between 8.1 mg and 15.1 mg among European women of reproductive age in a way not to exceed the total long-term daily iron intake from all sources of 40 mg/day as recommended by The European Food Safety Authority (EFSA), thereby maintaining safety while addressing deficiency ^31,32^. Another important strength of the study is the inclusion of a ferrous sulfate arm. Ferrous sulfate, known for its high bioavailability, is the most commonly used oral iron formulation in both therapeutic and preventive settings, and it frequently serves as a benchmark for evaluating the efficacy of new interventions ^17^. Although the study was not designed for a direct comparative analysis between the two intervention arms, the inclusion of the FSC group nonetheless provides valuable context for interpreting the relative effectiveness of the YIFC formulation under identical experimental conditions.

## Conclusions

The findings of the study highlight the underrecognition of dietary ID in an at-risk population within a developed country, emphasizing the relevance of targeted screening programmes and/or sustained supplementation strategies to support public health. Proactive low-dose iron supplementation in the form of either *>Your< Iron Forte Capsules (YIFC)* or *Ferrous Sulfate Capsules (FSC)* was demonstrated to be a practical, well accepted, and well-tolerated approach to effectively address dietary iron gaps in otherwise healthy individuals with low iron stores, as the interventions significantly increased hemoglobin and serum ferritin levels, with the magnitude and the dynamics of changes within physiological levels and consistent with the population baseline iron status, the dose of iron provided, and the duration of the intervention. Both supplements were well tolerated, with *YIFC* being slightly advantageous over *FSC* in that adverse events with a definite attribution to the study product were significantly less frequent in the *YIFC* arm and in contrast to *FSC*, limited to the gastrointestinal system only. The dosing regimen and the nature of both supplements were compatible with everyday lifestyle habits of the study population, suggesting strong potential for long-term adherence and successful integration into routine use.

## Methods

### Study Design

This randomized, double-blind, parallel-design, single-center study evaluated 12 weeks of once-daily iron supplementation in adults with nutritional iron deficiency. The study was conducted between January 2022, and January 2024, in Ljubljana, Slovenia. The study adhered to the International Conference on Harmonization Good Clinical Practice guidelines, the Declaration of Helsinki, and relevant clinical research regulations and guidelines. Ethical approval was granted by the Nutritional Research Ethics Committee at Biotechnical Faculty, University of Ljubljana. The study was registered at Clinicaltrials.gov (NCT05185024; https://clinicaltrials.gov/study/NCT05185024) on December 20^th^, 2021. Although the public was not directly involved in the design, the conduct, the reporting, or the dissemination plans of this study, feedback from participants during the early phases of the study informed protocol amendments ensuring better alignment with their neds and contributed positively to the study’s execution.

### Participants

Study participants were recruited from the public via influencer marketing and outreach at a healthy lifestyle fair and several faculties in Ljubljana between January 2021 and April 2023. Participants were directed to a single healthcare facility, Medicare Plus, d. o. o., which was used as the only study site. Participants were required to meet the following inclusion criteria: personally signed and dated informed consent form (ICF) (separate for screening and for main study); male or female, aged 18-50 years (inclusive); mild to moderate iron deficiency (hemoglobin (Hb) value of 100-130 g/L (women) and 110-140 g/L (men) and a serum ferritin (SF) level <30 µg/L (with concomitant C-reactive protein (CRP) level <10 mg/L)); generally healthy with no known significant health problems as judged based on medical history, the results of clinical laboratory tests performed at screening, and the results of examination of vital signs; Body Mass Index (BMI) <27 kg/m^2^; and ability to understand and willingness to comply with all study procedures.

Exclusion criteria included: gastrointestinal bleeding (as determined by the fecal occult blood test (FOBT)); pronounced symptoms accompanying ID; hemochromatosis or other iron-loading disorders; known hemoglobinopathy; any active or unstable concurrent medical condition; presence of an inflammatory and/or autoimmune disorder; partial or total gastrectomy or any other surgical procedure bypassing the duodenum; known human immunodeficiency virus (HIV), hepatitis B virus (HBV), or hepatitis C virus (HCV) infection; any active chronic or acute infectious disease requiring antibiotic treatment; taking regular medication that could affect iron absorption; use of any iron-containing medications or supplements within 30 days prior to screening and at any time during the study; receipt of blood transfusion within 12 weeks prior to screening or planned blood transfusion during the study; receipt of intramuscular (IM) or intravenous (IV) injection of depot iron preparation within 30 days prior to screening or at any time during the study; receipt of erythropoiesis-stimulating agents within 30 days prior to screening or at any time during the study; blood donation within the previous 30 days prior to screening or at any time during the study; scheduled or expected hospitalization and/or surgery during the course of the study; intake of concurrent medications that could interfere with physical or mental performance; for women of childbearing potential: planning of pregnancy, current pregnancy, or lactation; known allergies or severe adverse reactions to previous oral iron therapy or known hypersensitivity to any component of investigational products; current participation in any other interventional clinical study or participation within 30 days prior to screening; any other unspecified reason which, in the opinion of the investigator, made the individual unsuitable for participation.

### Investigational Products

The investigational products (IPs) used in this study were *>Your< Iron Forte Capsules (YIFC)* and *Ferrous Sulfate Capsules (FSC)*, containing 30 mg of elemental iron in the form of Qfer® and ferrous sulfate, respectively, and 60 mg of vitamin C. To achieve blinding, both products were matched in appearance (size, shape, color), smell, and packaging. Both products were developed and supplied by PharmaLinea Ltd. (Ljubljana, Slovenia, EU) and manufactured under FSSC 22000 and Good Manufacturing Practice for Food Supplements. Both products were quality tested by independent external accredited laboratories.

Randomization to an investigational product was performed in a 1:1 ratio by the investigator according to a pre-defined randomization schedule created with Microsoft Excel 2019 (Microsoft, Redmond, WA, USA) by the study statisticians, using a permuted block randomization approach with a fixed block size of 12. Participants as well as the investigator remained blinded to IP allocation throughout the duration of the study. Efficacy analysis and analysis of harms after study completion were performed on blinded data sets and IP identity of each data set was only revealed when the analyses were completed.

IPs were labelled with a unique product code based on the pre-specified randomization list by the study manager not involved in the conduct of the study. Each participant was assigned sets of IPs with the same product code for the whole duration of the study. The daily dosing of the IPs for each participant consisted of one capsule to be ingested with a glass of water on an empty stomach, separated from intake of food, coffee, tea, or milk by at least one hour. Compliance with the consumption of the IPs was assessed by counting the number of returned capsules at each relevant study visit. Adequate IP compliance was defined as 80%-120% compliance with the dosage schedule.

### Study Procedures

Individuals interested in participating were first scheduled to undergo a screening procedure to determine their potential eligibility for participation in the study. Upon signing of informed consent for screening, a unique screening identified was assigned to each participant and a questionnaire on general health-, lifestyle- and diet-related information, including questions on selected inclusion and exclusion criteria, was administered to determine their eligibility. Hemoglobin was measured using a point-of-care finger prick test to pre-select individuals with values within the target ranges (100–130 g/L (women), 110–140 g/L (men)). Eligible candidates then provided a fasted morning venous blood sample to assess the level of SF (in combination with CRP) and to obtain baseline hematological data. Participants meeting criteria underwent FOBT to exclude gastrointestinal bleeding; individuals with negative test results were invited to participate in the study. All screening procedures were completed within 21 days of consent.

The study consisted of 12 weeks of once-daily dosing with IPs. During this period, three in-person study visits were conducted. Enrollment into the study occurred at the first study visit after signing of informed consent for participation in the main study. Each enrolled participant was assigned a unique study identifier in the form of an extension to their screening identifier. Eligibility against all remaining inclusion and exclusion criteria was evaluated, vital signs checked, basic demographic information collected, important medical history recorded, information on concomitant medications and therapies gathered, and enquiry on any adverse events (AEs) since screening made. Eligible participants were randomized to receive either YIFC or FSC. At each of the first two visits, participants received a supply of IPs to last until their next study visit, and a study diary to record any missed IP doses, AEs and other important health- or lifestyle-related information occurring in the periods between the study visits. At the second and the third study visits, scheduled for four and 12 weeks after randomization, respectively, a fasted morning venous blood sample was drawn from each participant to evaluate relevant hematological (Hb, red blood cell (RBC) indices), iron profile (serum iron, SF, total iron-binding capacity of transferrin (TIBC), transferrin saturation (TSAT)), and safety/eligibility (beta-human chorionic gonadotropin (beta hCG)) parameters. Vital signs and demographic information were checked, information on concomitant medications and therapies gathered, and enquiry on any AEs occurring since previous study visit made. Any participant presenting with an ongoing AE at the third study visit or at the time of their premature removal from the study was followed-up for up to four weeks for AE resolution.

### Objectives and Outcomes

The primary outcome was the change from baseline in Hb after 12 weeks of supplementation with either IP. Secondary outcomes included changes in Hb after four weeks of intervention and changes in RBC indices (hematocrit (Hct), MCV (Mean Corpuscular Cell Volume), MCH (Mean Corpuscular Hb), MCHC (Mean Corpuscular Hb Concentration)) and iron-related biomarkers (SF, TSAT) at both four and 12 weeks of intervention. The difference in Hb increase between the two study arms after 12 weeks of intervention was also set as a secondary objective.

Safety objectives included assessment of the incidence and nature of AEs over 12 weeks. AEs were collected non-systematically via participant diaries and investigator inquiries and were assessed by the investigator for seriousness, severity, attribution, and outcome.

An exploratory objective was the evaluation of fatigue using self-administered pen-and-paper format of the Energy/fatigue subscale of the RAND 36-Item Health Survey (version 1.0), administered at weeks four and 12 post-intervention start and scored per RAND guidelines (www.rand.org).

### Sample Size and Statistical Analyses

Sample size calculations were based on the primary outcome, a change in Hb level between baseline and 12 weeks of IP intervention. Assuming a normal distribution with a standard deviation (SD) of 8 g/L ^33^ and a true difference of 6 g/L in the mean Hb level of matched pairs, 44 participants per arm were calculated to be required to achieve 90% power at a 5% significance level. Accounting for an anticipated 15% dropout rate, the sample was increased by 6 individuals per arm.

Statistical analyses were conducted using SPSS Statistics for Windows, Version 29.0 (IBM Corp., Armonk, NY, USA) and Microsoft Office Excel 2016 (Microsoft, Redmond, WA, USA). Three populations were defined for efficacy analysis: the intention-to-treat (ITT) population included all randomized participants, regardless of whether they completed the study or fully adhered to the protocol; the full analysis set (FAS) population included all randomized participants for whom a baseline measurement as well as the measurement at 12 weeks post-randomization were available, regardless of their protocol adherence; the per-protocol (PP) population included a subset of the participants of the FAS population – those compliant with IP consumption and without major protocol deviations in terms of prohibited concomitant therapies. Efficacy analyses were conducted in all three populations; FAS results are presented when findings were consistent across populations. Primary and secondary outcomes were analyzed per each study arm using a two-sided paired t-test (or equivalent non-parametric test in case the data were not normally distributed). For the primary outcome, one pre-defined sub-group analysis was performed, stratifying participants according to their baseline Hb into either low (<120 g/L for women and <130 g/L for men) or normal (≥120 g/L for women and ≥130 g/L for men) baseline Hb.

Differences between these subgroups were analyzed for each study arm separately, using a two-sided paired t-test (or equivalent non-parametric test in case the data were not normally distributed), as well as between the study arms, using a t-test for independent samples (or equivalent non-parametric test in case the data were not normally distributed). A non-inferiority analysis comparing Hb levels at 12 weeks of intervention between YIFC and FSC was performed applying a non-inferiority margin of 4 g/L for the lower bound of the 95% confidence interval.

Analysis of harms was performed on the safety population (for characterization of the safety population refer to **Figure 1** and **Adverse Events** section). Adverse event frequencies and the frequencies of participants experiencing AEs were compared between study arms using Fisher’s exact probability and Chi-square tests. Adverse events were coded with Medical Dictionary for Regulatory Activities (MedDRA). Characteristics of AEs (seriousness, severity, attribution, outcome) were reported descriptively.

For the exploratory outcome, changes in the Energy/fatigue subscale of the RAND 36-Item Health Survey were analyzed using two-sided paired t-tests for each arm to evaluate the changes at weeks four and 12.

## Acknowledgements

The authors wish to thank the study participants and acknowledge the support of personnel involved in the conduct of the study: Ms. Bianca Bevc (head study nurse; Medicare Plus d. o. o.), Ms. Nives Vivod (study nurse; PharmaLinea Ltd.), Mr. Aleš Koščak (study technician; Medicare Plus d. o. o.), Ms. Barbara Ježek (study nurse; Medicare Plus d. o. o.), Ms. Sanja Milijaš (study administrator; Medicare Plus d. o. o.), prim. Jožef Pretnar, MD, spec. internal med. and hematology (medical advisor), Ms. Ana Vizler (coordination of screening, data entry).

## Author Contributions

Conceptualization, methodology, visualization, G.P.; investigation, A.K., B.D., N.F.; data curation and formal analysis, I.L., N.Č.L., N.R.M., M.K.; writing—original draft, G.P., T.D.; writing—review and editing, A.K., B.D., N.F., I.L., N.Č.L., N.R.M., M.K., T.D., G.P. All authors have read and agreed to the published version of the manuscript.

## Data Availability

The materials that support the findings of this study are available from the corresponding author, G.P., upon reasonable request and with permission of PharmaLinea Ltd. Access to anonymised data is limited to the principal investigator, A.K.

## Additional information

### Supplementary Information

The online version contains supplementary material available at https://….

### Note to Readers

The study results are specifically applicable to *>Your< Iron Forte Capsules* and *Ferrous Sulfate Capsules* and should not be generalized to other similar products that use different formulations, such as syrups or sprays.

### Ethical approval and consent to participate

The study was approved by the Nutritional Research Ethics Committee at Biotechnical Faculty, University of Ljubljana (Ref. No. KEP-1-6/2021, approved November 22nd, 2021). Written informed consent was obtained from all participants prior to their inclusion in the study.

### Open Access

This article and supplementary material are open access.

## References

1. Camaschella, C. Iron-deficiency anemia. N Engl J Med 372, 1832–43 (2015).

2. Pasricha, S.-R., Tye-Din, J., Muckenthaler, M. U. & Swinkels, D. W. Iron deficiency. Lancet 397, 233–248 (2021).

3. Nutritional anaemias: tools for effective prevention and control. Geneva: World Health Organization; 2017. Licence: CC BY-NC-SA 3.0 IGO.

4. Iolascon, A. et al. Recommendations for diagnosis, treatment, and prevention of iron deficiency and iron deficiency anemia. Hemasphere 8, e108 (2024).

5. Milman, N. T. Dietary Iron Intake in Women of Reproductive Age in Europe: A Review of 49 Studies from 29 Countries in the Period 1993-2015. J Nutr Metab 2019, 7631306 (2019).

6. Milman, N., Taylor, C. L., Merkel, J. & Brannon, P. M. Iron status in pregnant women and women of reproductive age in Europe. Am J Clin Nutr 106, 1655S–1662S (2017).

7. Lavriša, Ž. et al. Dietary Iron Intake and Biomarkers of Iron Status in Slovenian Population: Results of SI.Menu/Nutrihealth Study. Nutrients 14, (2022).

8. Camaschella, C. & Girelli, D. The changing landscape of iron deficiency. Mol Aspects Med 75, 100861 (2020).

9. Patterson, A. J., Brown, W. J., Roberts, D. C. & Seldon, M. R. Dietary treatment of iron deficiency in women of childbearing age. Am J Clin Nutr 74, 650–6 (2001).

10. Garcia, O. P., Diaz, M., Rosado, J. L. & Allen, L. H. Ascorbic acid from lime juice does not improve the iron status of iron-deficient women in rural Mexico. Am J Clin Nutr 78, 267–73 (2003).

11. Kantor, E. D., Rehm, C. D., Du, M., White, E. & Giovannucci, E. L. Trends in Dietary Supplement Use Among US Adults From 1999-2012. JAMA 316, 1464–1474 (2016).

12. Papatesta, E. M., Kanellou, A., Peppa, E. & Trichopoulou, A. Is Dietary (Food) Supplement Intake Reported in European National Nutrition Surveys? Nutrients 15, (2023).

13. Liu, L., Tao, H., Xu, J., Liu, L. & Nahata, M. C. Quantity, Duration, Adherence, and Reasons for Dietary Supplement Use among Adults: Results from NHANES 2011-2018. Nutrients 16, (2024).

14. Norman, G. R., Sloan, J. A. & Wyrwich, K. W. Interpretation of changes in health-related quality of life: the remarkable universality of half a standard deviation. Med Care 41, 582–92 (2003).

15. WHO guideline on use of ferritin concentrations to assess iron status in individuals and populations. Geneva, Switzerland: World Health Organization; 2020. Licence: CC BY-NC-SA 3.0 IGO. Available at https://www.who.int/publications/i/item/9789240000124.

16. Hallberg, L. et al. Screening for iron deficiency: an analysis based on bone-marrow examinations and serum ferritin determinations in a population sample of women. Br J Haematol 85, 787–98 (1993).

17. Low, M. S. Y., Speedy, J., Styles, C. E., De-Regil, L. M. & Pasricha, S.-R. Daily iron supplementation for improving anaemia, iron status and health in menstruating women. Cochrane Database Syst Rev 4, CD009747 (2016).

18. Casgrain, A., Collings, R., Harvey, L. J., Hooper, L. & Fairweather-Tait, S. J. Effect of iron intake on iron status: a systematic review and meta-analysis of randomized controlled trials. Am J Clin Nutr 96, 768–80 (2012).

19. Simic, S. et al. Effectiveness of low-dose iron treatment in non-anaemic iron-deficient women: a prospective open-label single-arm trial. Swiss Med Wkly 153, 40079 (2023).

20. Stefan, M. W. et al. Assessment of the Efficacy of a Low-Dose Iron Supplement in Restoring Iron Levels to Normal Range among Healthy Premenopausal Women with Iron Deficiency without Anemia. Nutrients 15, (2023).

21. D’Adamo, C. R., Novick, J. S., Feinberg, T. M., Dawson, V. J. & Miller, L. E. A Food-Derived Dietary Supplement Containing a Low Dose of Iron Improved Markers of Iron Status Among Nonanemic Iron-Deficient Women. J Am Coll Nutr 37, 342–349 (2018).

22. Soppi, E. Iron Deficiency in the Absence of Anemia - A Common, Complex and Challenging Disease to Treat. Med Res Arch (2022) doi:10.18103/mra.v10i10.3224.

23. Moretti, D. et al. Oral iron supplements increase hepcidin and decrease iron absorption from daily or twice-daily doses in iron-depleted young women. Blood 126, 1981–9 (2015).

24. Stoffel, N. U. et al. Iron absorption from oral iron supplements given on consecutive versus alternate days and as single morning doses versus twice-daily split dosing in iron-depleted women: two open-label, randomised controlled trials. Lancet Haematol 4, e524–e533 (2017).

25. Tolkien, Z., Stecher, L., Mander, A. P., Pereira, D. I. A. & Powell, J. J. Ferrous sulfate supplementation causes significant gastrointestinal side-effects in adults: a systematic review and meta-analysis. PLoS One 10, e0117383 (2015).

26. Stoffel, N. U., von Siebenthal, H. K., Moretti, D. & Zimmermann, M. B. Oral iron supplementation in iron-deficient women: How much and how often? Mol Aspects Med 75, 100865 (2020).

27. Snook, J. et al. British Society of Gastroenterology guidelines for the management of iron deficiency anaemia in adults. Gut 70, 2030–2051 (2021).

28. Dugan, C. et al. The misogyny of iron deficiency. Anaesthesia 76 Suppl 4, 56–62 (2021).

29. Houston, B. L. et al. Efficacy of iron supplementation on fatigue and physical capacity in non-anaemic iron-deficient adults: a systematic review of randomised controlled trials. BMJ Open 8, e019240 (2018).

30. Yokoi, K. & Konomi, A. Iron deficiency without anaemia is a potential cause of fatigue: meta-analyses of randomised controlled trials and cross-sectional studies. Br J Nutr 117, 1422–1431 (2017).

31. Rippin, H. L., Hutchinson, J., Jewell, J., Breda, J. J. & Cade, J. E. Adult Nutrient Intakes from Current National Dietary Surveys of European Populations. Nutrients 9, (2017).

32. EFSA Panel on Nutrition, N. F. and F. A. (NDA) et al. Scientific opinion on the tolerable upper intake level for iron. EFSA J 22, e8819 (2024).

33. Ahmed, F., Khan, M. R. & Jackson, A. A. Concomitant supplemental vitamin A enhances the response to weekly supplemental iron and folic acid in anemic teenagers in urban Bangladesh. Am J Clin Nutr 74, 108–15 (2001).

