## Supplementary Table 1 for "The Efficacy and Safety of Daily Low-Dose Iron Supplementation in Adults with Dietary Iron Deficiency: a Double-Blind, Randomized, Single-Center Study"

**Supplementary material**

### Adverse Events

Of 334 AEs reported by 85 participants, data for four AEs reported by four participants were either missing or incomplete. Two of these four participants acknowledged experiencing an AE but declined to provide any details, preventing AE classification or categorization. The other two AEs were assigned to the MedDRA System Organ Classes (SOCs) of Injury, poisoning and procedural complications (knee injury) and Reproductive system and breast disorders (worsening of endometriosis requiring surgery, also classified as a serious adverse event (SAE)). However, data on severity, attribution, and outcome were missing for both, rendering them non-evaluable. As a result, four participants and four AEs had to be excluded from the AE analysis, leaving 81 evaluable participants experiencing 330 evaluable AEs in the safety analysis set.
Of the 330 evaluable AEs reported, one was classified as a SAE under the Gastrointestinal disorders MedDRA SOC. The participant experiencing this SAE was hospitalized due to nausea and vomiting. The SAE was graded as severe but not related or not likely to be related to the IP and the participant went on to complete the study. Three hundred twenty-seven AEs were assessed as mild or moderate, and three as severe. None of the severe AEs were considered causally related to the IP. The vast majority of AEs (328 events, 99%) did not require any action to be taken regarding the study, however, two AEs (1%) occurring in two separate participants led to study discontinuation. One case presented as an unspecified itchy rash (moderate, probably related to the IP) and the other was a deep cut to the thumb requiring systemic antibiotic treatment (moderate, IP non-related).
By the end of the study, 319 AEs (97%) had resolved with full recovery. Two participants were still recovering from acute nasopharyngitis (both IP non-related). Another three participants had a total of six ongoing AEs: three were gastrointestinal disorders (constipation, functional dyspepsia, and abdominal distension) judged as IP-related, and the other two were amenorrhea and patellar tendonitis – IP non-related. The statuses of six AEs in three participants remained unknown due to lack of follow-up information.
Overall, no safety concerns were identified in association with the consumption of either of the IPs. There were no significant differences in the number of participants experiencing AEs or the frequency of AEs between the study arms, with the exception of AEs classified as definitely related to the IP, which occurred significantly less often in the YIFC arm.

| Parameter | Study population* by intervention arm | | ALL (N=97) |
|---|---|---|---|
| | YIFC (N=51) | FSC (N=46) | |
| **AEs,** N | | | |
| AEs | 177 | 153 | 330 |
| SAEs | 1 | 0 | 1 |
| **Participants**[a], N (%)[b] | | | |
| Participants with AEs | 43 (84) | 38 (83) | 81 (84) |
| Participants with SAEs | 1 (2) | 0 (0) | 1 (1) |
| **AEs by severity**, N (%)[c] | | | |
| Mild | 65 (37) | 77 (50) | 142 (43) |
| Moderate | 109 (62) | 76 (50) | 185 (56) |
| Severe | 3 (2) | 0 (0) | 3 (1) |
| **Participants with AEs by severity**[a], N (%)[b] | | | |
| Mild | 26 (60) | 32 (84) | 58 (72) |
| Moderate | 36 (84) | 29 (76) | 65 (80) |
| Severe | 3 (7) | 0 (0) | 3 (4) |
| **AEs by causality**, N (%)[c] | | | |
| Not related | 105 (59) | 88 (58) | 193 (59) |
| Not likely | 46 (26) | 35 (23) | 81 (25) |
| Possibly | 20 (11) | 16 (10) | 36 (11) |
| Probably | 4 (2) | 6 (4) | 10 (3) |
| Definitely[†] | 2 (1) | 8 (5) | 10 (3) |
| **Participants with AEs by causality**[a], N (%)[b] | | | |
| Not related | 37 (86) | 32 (84) | 69 (85) |
| Not likely | 16 (37) | 16 (42) | 32 (40) |
| Possibly | 12 (28) | 11 (29) | 23 (28) |
| Probably | 3 (7) | 5 (13) | 8 (10) |
| Definitely | 1 (2) | 4 (11) | 5 (6) |
| **AEs by action taken**, N (%)[c] | | | |
| No action | 176 (99) | 152 (99) | 328 (99) |
| Study discontinuation | 1 (1) | 1 (1) | 2 (1) |
| **Participants with AEs by action taken**[a], N (%)[b] | | | |
| No action | 42 (98) | 37 (97) | 79 (98) |
| Study discontinuation | 1 (2) | 1 (3) | 2 (2) |
| **AEs by outcome**, N (%)[c] | | | |
| Full recovery | 172 (97) | 147 (96) | 319 (97) |
| Recovering | 1 (1) | 1 (1) | 2 (1) |
| Ongoing | 1 (1) | 2 (1) | 3 (1) |
| Unknown | 3 (2) | 3 (2) | 6 (2) |
| **Participants with AEs by outcome**[a], N (%)[b] | | | |
| Full recovery | 41 (95) | 37 (97) | 78 (96) |
| Recovering | 1 (2) | 1 (3) | 2 (2) |
| Ongoing | 1 (2) | 2 (5) | 3 (4) |
| Unknown | 2 (5) | 1 (3) | 3 (4) |

**Supplementary Table 1.** Summary of all adverse events by study intervention arm. * Safety population. [a] A single participant may experience more than one AE with different severity or attribution grades, actions taken, and AE outcomes. [b] Percentages are based on the total number of participants with AEs in each study arm or cumulatively. [c] Percentages are based on the total number of AEs reported in each study arm or cumulatively. YIFC: >Your< Iron Forte Capsules, FSC: Ferrous Sulfate Capsules. AE: adverse event, SAE: serious adverse event. [†] Statistically significantly different at $p<0.05$.
